# Analysis of Factors Determining Success in FFPE Based NGS Panel Testing For Lung & Ovarian Cancer

**DOI:** 10.1101/2024.07.29.24311158

**Authors:** P. Bhattacharya, P. Bishop, D. Gokhale, S. Rowlston, B. Eggington, G. J. Burghel, H. Schlecht

**Affiliations:** Manchester Centre for Genomic Medicine, Manchester University Foundation Trust, Manchester, United Kingdom; Department of Pathology, Manchester University Foundation Trust, Manchester, United Kingdom; Division of Evolution, Infection and Genomic Sciences, School of Biological Sciences, Faculty of Biology, Medicine and Health, The University of Manchester, Manchester, UK

**Keywords:** NGS sequencing, Lung Cancer, Ovarian Cancer

## Abstract

The identification of oncogenic variants in both lung and ovarian cancer is central to personalised treatment. NGS approaches using formalin-fixed paraffin-embedded (FFPE) tissue samples are routinely implemented for variant detection, but optimisation of pre-analytic factors is critical for success. We performed a large multi-cohort retrospective audit assessing pre-analytic factors related to Qiagen in house custom designed NGS panel testing for lung (n=801 from 23 referring hospitals) and ovarian (n=882 from 85 referring hospitals) FFPE cancer samples, sequenced at the NHS Northwest Genomic Laboratory Hub (NWGLH). A further detailed analysis of a cohort of lung samples (n=461) submitted from a single high-volume referral centre was also undertaken. Overall NGS cohort success ranged from 74-85% with large variation amongst referring laboratories. Multivariate logistic regression analysis revealed DNA yield and quality to be significant predictors of NGS success (p<0.001) alongside sample type for lung (p=0.035) and use of macrodissection for ovarian (p=0.025). Univariate analysis revealed specific poor lung performance within biopsy samples and associated with number and length of core biopsy samples. Furthermore, excessive fixation time for lung cytology and ovarian samples was associated with NGS failure (p<0.05), with only 49.5% of lung endoscopic bronchial ultrasound (EBUS) cytology samples meeting existing local guidelines for fixation time <24 hours, with 2/3 of prolonged fixation samples being received in the lab the following day. Variation in key identified pre-analytic factors amongst referring centres and variable adherence to best practice guidelines is likely to be responsible for the wide variation in NGS success. Improved collaboration between NHS genomic hubs and referring pathology laboratories through the establishment of a regional interactive collaborative network, facilitating guideline sharing and assessing adherence to pre-analytic optimisation of sample collection and processing for genomic testing, is crucial to improve future NGS genomic cancer testing.

## Introduction

In recent years, the identification of targetable genetic aberrations in lung cancer, particularly non-small cell lung cancer (NSCLC), has revolutionized clinical practice (1). The development and implementation of routine molecular testing have been greatly facilitated by advances in next-generation sequencing (NGS) technology (2). NSCLC exhibits a wide range of molecular aberrations, including somatic variants, copy number aberrations, chromosomal rearrangements, and epigenetic dysregulation (3–5).

To diagnose lung cancer, a combination of detailed radiological imaging techniques such as computed tomography (CT), magnetic resonance imaging (MRI), and positron emission tomography (PET) is utilized (6). Invasive procedures, both internal (bronchoscopy, endobronchial ultrasound-guided fine needle aspiration [EBUS-FNA], endoscopic ultrasound-guided fine needle aspiration [EUS-FNA]) and external (CT-guided or ultrasound-guided percutaneous fine needle aspiration or core biopsy), are also employed to obtain pathological samples for diagnosis (6).

Since the majority of NSCLC patients present with advanced disease, it is recommended to make diagnoses using small biopsy or cytology-type samples, with the option to supplement standard histopathology with immunohistochemistry (IHC) to facilitate accurate diagnosis (6). The National Institute for Health and Care Excellence (NICE) guidelines for lung cancer play a crucial role in determining specific diagnostic pathways within the National Health Service (NHS), and decisions are often made through multidisciplinary team meetings (7).

Among the minimally invasive methods used for sample collection, endobronchial ultrasound-guided fine needle aspiration (EBUS-FNA) has gained importance. It allows for cytological and histological tissue collection, particularly from mediastinal lymph nodes, with real-time imaging guidance (8). The NICE guidelines emphasize the significance of EBUS-FNA, stating that regional cancer centres should have the capability to offer timely access to this technique, with regular audits of its adequacy (7).

Routine genetic biomarker analysis is now performed within the NHS, as targeted biological therapies have proven effective in such cases (9). After establishing a diagnosis, molecular diagnostic techniques are employed to detect targetable oncogenic alterations or provide information on immuno-oncology therapy biomarkers for individual patients. The focus is primarily on the most common oncogenic driver genes observed in NSCLC, including *EGFR, ALK, ROS1, BRAF, NTRK, HER2,* and fusion genes involving *RET* and *MET* exon 14 variants (6).

NGS has become a widely used method for variant detection due to its numerous advantages, such as the ability to detect both small variants and larger structural rearrangements, the short time required for library preparation, and the relatively low cost (10). Compared to standard polymerase chain reaction (PCR) testing, NGS offers higher sensitivity and specificity while assessing a range of variants from a relatively small amount of sample (10).

The successful implementation of NGS technologies relies on optimal processing and preservation of specimens, especially small diagnostic samples, in referring diagnostic pathology laboratories. These factors significantly influence subsequent successful genomic analysis (11). While guidelines often focus on reporting, bioinformatics, and analytical wet bench processes, it is equally important to consider pre-analytic factors related to the acquisition and processing of samples (11).

Factors that impact NGS success include DNA quantity, DNA quality, specimen type, and tumour cellularity (12). Formalin fixation is a standard method for preserving biopsy and surgical excision samples, with subsequent embedding in paraffin wax blocks (FFPE). However, formalin fixation can cause fragmentation of nucleic acids and random DNA breaks, which can affect downstream analyses (13). For cytology specimens, the fixative CytoRich Red has been shown to be superior to formalin in maintaining DNA quality (14).

The success of NGS also depends on the yield and amount of DNA input required for the specific NGS platform used (12). The size of the tumour-rich area within a specimen and the number of viable tumour cells obtained are critical factors influencing NGS success (15). Additionally, the presence of necrosis within a specimen can impact the reliability of NGS results (16).

The type of tumour specimen is also a crucial consideration. Fine needle aspirations and core biopsies, although minimally invasive and often used for anatomically difficult lesions, generally provide a smaller amount of tissue, while surgical excisions yield greater amounts of DNA (15). The neoplastic cell count, or tumour fraction, is another factor influencing NGS success in identifying relevant genetic variants when present {Roy-Chowdhuri, 2015 #53;Burghel, 2019 #41}. In cytological specimens, the choice of glass slide used for sample preparation can affect DNA quantities, with frosted tip slides yielding higher DNA quantities (18).

Ovarian cancer, like other cancers, exhibits a wide range of genomic variations, including *TP53* variants, copy number gains in genes such as *BRAF, TERT, TERC,* and *CCNE1*, germline *BRCA1/2* variants and copy number loss of *RB1* with or without *PTEN* (19–21). The NHS genomics testing strategy for ovarian cancer focuses on associated somatic small variants, structural rearrangements, and constitutional testing (22). Homologous recombination deficiency (HRD) status and *NTRK* fusion genes are important considerations for treatment decisions (23). Constitutional testing is performed for individuals diagnosed with high-grade serous ovarian cancer using an NGS multigene panel (22).

Differences in sample processing and sequencing may explain the higher success rates observed in ovarian cancer compared to lung cancer assays. Ovarian cancer assays have success rates ranging from approximately 96% to 100%, while lung cancer assays range from 69% to 91% (24, 25). This difference can be attributed to the greater use of surgical excision specimens for NGS in ovarian cancer, resulting in higher cellular content for DNA extraction. Validation studies for NGS in ovarian cancer often utilize samples with higher nuclear cell content (>30%), compared to the >20% typically employed in lung cancer studies (26).

The need for standardized protocols to maximize NGS success for ovarian cancer samples has been highlighted in previous studies. Various pre-analytic factors, including tissue cold ischemic times, minimum and maximum fixation times, and sample storage and processing, can affect molecular integrity and NGS results (12, 17). Other key pre-analytic factors that may potentially influence NGS success are tissue to fixative ratio, fixative concentration, slide drying duration, and temperature, but they still lack sufficient evidence in the literature to make specific recommendations. The aim of this study was to use large locally available datasets from samples processed at the Northwest Genomic Laboratory Hub (NWGLH) sent from various regional, national, and international centres to assess adherence to previously published regional NGS guidelines (11, 27) and provide new recommendations to improve success rates of NGS multi-panel testing.

## Materials and Methods

### Selection of Test Data Groups

A retrospective audit was conducted to assess pre-analytic factors in three cohorts of lung and ovarian cancer samples analysed using a custom next-generation sequencing (NGS) panel at Manchester Foundation Trust (MFT). Patient demographic data, including age, gender, final diagnosis, and referring hospital, were extracted for all patients included in the study. The following pre-analytic variables were recorded from available genetic or pathology databases for further analysis: DNA concentration (obtained from genomics lab quality control data and measured in ng/µl), DNA quality (obtained from genomics lab quality control data and assessed using Nanodrop 230/280 ratio and 260/280 ratio), Presence of macrodissection (extracted from pathologists’ comments indicating whether macrodissection was performed), Neoplastic cell content (extracted from pathologists’ comments and categorized into three groups based on documented subjective visual assessment: <20%, >20%, or >50% neoplastic cells), Sample Type (broadly divided into surgical excision and biopsy samples. Biopsy samples included core biopsies and fine needle aspiration samples with cytology. For lung cancer samples, the methods of biopsy acquisition were further analysed, particularly focusing on radiologically guided (US/CT), bronchoscopic, or endobronchial ultrasound (EBUS) guided procedures. Anatomical location, such as lymph node station and position, was also recorded for EBUS samples, Tissue fixation time (recorded as “days to receive” and “days to process” where data were available).

The initial observational approach aimed to identify high-volume referrers and sample types with the lowest performance for subsequent detailed analysis. By comparing two different cancer types, the study aimed to identify common pre-analytic factors that are crucial and to identify specific factors that could be improved.

### Study sample size/power calculation

Following consultation with a University of Manchester biostatistician, a power calculation (using clinicalc.com (79)) was performed to assess optimal sample size for the study. Considering a dichotomous endpoint with two independent samples as the basis of statistical testing and a power of 80%, α= 0.05, an estimated 10% difference between the two testing groups (85% v 95%) with a 5 fold excess in terms of samples within the successful group based on a small pilot dataset of 50 samples, the optimal sample size was calculated at n=432.

### Sample Cohorts

To conduct the analysis, data from lung cancer samples processed over a six-month period (September 23, 2021, to March 23, 2022) was extracted with the help of the bioinformatics team at the Manchester Genomics Centre. Analytic and pre-analytic data were collected for each individual sample, and incomplete datasets were excluded. The final cohort consisted of 801 samples for further association assessment and statistical analysis.

Additionally, in collaboration with key collaborators at Wythenshawe Hospital, more detailed pre-analytic information, including fixation time, was extracted from their cohort of referred lung cancer samples over a 17-month period (January 1, 2021, to June 1, 2022). This produced a separate cohort of 461 samples, which was subjected to further detailed analysis. Fixation time data was available for biopsy samples only within this cohort.

Similarly, an ovarian cancer cohort processed over a seventeen-month period (January 1, 2021, to June 1, 2022) at MFT was included in the analysis. After exclusions, the final cohort consisted of 882 samples. Within this dataset, additional information was gathered for 30 samples referred from MFT itself and 29 samples from the Royal Preston Hospital.

### Pre-analytic Pathology lab sample processing

The key steps of the process required to produce FFPE histopathology sections that are then used for DNA are: Fresh tissue specimen acquisition, Fixation, Dehydration, Clearing, Wax infiltration and Embedding. The steps and tissue fixation methods used for FFPE sample preparation from surgical excision specimens and core biopsies, differ from those required for cytology specimens. Different referring laboratories also use slightly different methods based on locally agreed protocols for the various steps of the process.

All samples meeting essential acceptance criteria are subsequently processed. All Endobronchial biopsies / lung cores are prioritised as urgent. Existing guidelines for optimising the FFPE processing pathway for genomic samples are as follows-10% Neutral Buffered Formalin (4% formaldehyde) is recommended for fixation. This should be freshly diluted from stock ideally within the previous 24hrs. (According to Wythenshawe hospital protocols CytoRich Red (CRR) is used as a fixative for EBUS FNA and other FNA specimens to prepare cytology from).

Fixation should neither be too long or too short (not less than 6 hours, no more than 24 hours). Formalin fixation times longer than 24 hours should only be used when the characteristics of the specimen demand it (i.e. particularly large specimens).

Alternatives to pre-filled, pre-mixed formalin pots >24 hours old, although widely used, should be considered for genomic samples and emphasis should be placed on rapid transporting and pre-analytic processing of samples in local genomics laboratories.

For biopsy samples if more than one core is available, process identifiable cores into more than one block with remaining smaller fragments being incorporated into a single block; whereas, for cytology samples, where feasible, incorporate material maximally within a cell block preparation.

### DNA extraction from FFPE samples

Tissue clean-up and deparaffinization: Ethanol and xylene were used to remove impurities and deparaffinize the samples.

Cobas extraction kit method: Proteinase K reagent and DNA Tissue Lysis Buffer were added, followed by incubation and centrifugation steps.

DNA binding and washing: DNA Binding Buffer and Wash Buffer I were added to the samples, and centrifugation was performed to remove impurities.

DNA elution: DNA elution buffer was added, and the samples were centrifuged and transferred to labelled tubes.

DNA quantification and assessment: The Qubit dsDNA BR Assay Kit 2.0 Fluorometer and Nanodrop® 8000 spectrophotometer were used to quantify and analyse the DNA. In terms of DNA quality assessment, absorbance ratios of 260/280 (>1.8) and 260/230 (1.8-2.2) were monitored.

### NGS sequencing using QIAseq enrichment

NWGLH utilizes QIAseq enrichment methodology (developed by Qiagen) for NGS sequencing of NSCLC and ovarian cancer samples prior to NextSeq Illumina NGS sequencing. This approach incorporates molecular barcodes to enhance variant detection accuracy. QIASeq is a hybrid technology combining amplicon and sequence capture methods. Initially, 10-80 ng of fresh DNA or 100-250 ng of FFPE DNA is enzymatically fragmented and ligated with universal adaptors containing Unique Molecular Indexes (UMIs) at the 5’ ends.

### Bioinformatic analysis for variant identification

The QiaSeq LLM pipeline used in this study supports UMI counting. A minimum read depth of 138x UMIs is required for 95% confidence in capturing a variant at 4% variant allele frequency (VAF) on 2 or more UMIs, and 278x UMIs are needed for the same confidence at 2% VAF. MFT quotes variant detection to 4% VAF with a recommended minimum mean UMI read depth of 138x or 2% in hotspot regions. Variants can still be confidently called and reported if supported by more than one UMI, even if the mean UMI depth is below the recommended threshold. A sample with a mean UMI depth ≤50x should be failed, unless a true pathogenic variant is present. The procedure demonstrates 100% sensitivity, specificity, and reproducibility. Variants are checked on IGV using BAM files to assess coverage and strand-bias. Reliable variants are those found at reasonable frequency in reads from both strands.

The LLM pipeline is used for variant detection. Illumina NextSeq generates reads, which are aligned against the human genome using Stampy and BWA. VarScan, DREEP, and Pindel are employed for calling SNPs and indels. Variants are annotated to Ensembl, dbSNP, and OMIM databases. RefSeq transcripts are used for collapsing and filtering variants. Varscan variants with frequency <4% and strand bias are excluded. Variant calling is performed using a bed file region of interest. An excel report is generated containing SNP and indel calls, coverage graphs, and other relevant information.

A custom-designed amplicon-based QIAseq targeted DNA panel named CORE, comprising 37 genes of interest, is manufactured by Qiagen. All referred samples are sequenced for all panel genes, but only genes relevant to the clinical indication are analysed.

Hotspot variants with UMI supporting reads are reported without further interpretation at 2% VAF. Variants with >4% VAF require additional assessment based on ACGS/AMP guidelines. Pathogenic and likely pathogenic variants are recorded in the genotype box, while other somatic variants considered VUS/passengers are recorded in the comments field. Class 1 variants are not recorded. Upon completion of the analysis, a clinical report is issued, providing interpretation of all identified pathogenic or likely pathogenic variants.

### Statistical analysis (Multiple Logistic Regression, Univariate and Bivariate analyses)

A multivariate logistic regression was conducted to assess the relationship between Qiaseq Status and the explanatory variables (Concentration, Nanodrop 260 Ratio, Nanodrop 230 Ratio, and Sample Type) in the Large Lung NGS, Wythenshawe, and Ovarian datasets. Data were checked for multicollinearity using the Belsley-Kuh-Welsch technique. Heteroskedasticity and normality of residuals were evaluated using the White test and the Shapiro-Wilk test, respectively. A p-value < 0.05 was considered statistically significant, and patients with missing data were excluded from the analysis. Statistical analysis was performed using the online application EasyMedStat (version 3.21; www.easymedstat.com).

Further bivariate and univariate comparisons were made between Qiaseq status and Concentration, DNA quality, macrodissection, nuclear cell content, sample type, and fixation time. Normality and heteroskedasticity of the data were assessed using the Shapiro-Wilk test and Levene’s test. The association between a specific parameter and Qiaseq status was analysed using the Mann-Whitney test for continuous variables and Chi-squared test for discrete variables. Fischer’s exact test was performed for the ovarian cancer tissue fixation data due to the smaller sample size. An alpha risk of 5% (α = 0.05) was set, and statistical analysis was conducted using EasyMedStat (version 3.21; www.easymedstat.com).

## Results

### a) Overall NGS Panel Success Rates & Regional Variation

Considering Northwest lung samples, Analysis of the overall cohort of 801 lung NGS samples, processed over a 6-month time period from 23/09/21 to 23/03/22, revealed the overall number of samples within this cohort that were successfully sequenced by the MFT NGS custom panel to be 591, with 210 unsuccessful (overall success rate of 74%) (Table 1.). 23 different individual referring hospitals were identified from the sample cohort, with 136 samples (17%) classified as unknown from the available data, leaving n=665 for analysis. Whilst there were significant regional variations in the number of samples referred for sequencing, overall success rates also varied amongst individual referrers, ranging from 68% - 81% amongst the top five high-volume referrers. Furthermore, analysis of the overall cohort of 461 lung NGS samples specifically referred from Wythenshawe Hospital for NGS sequencing at the NWGLH, processed over a 17-month time period from 01/01/2021 to 01/06/2022, revealed the overall number of samples within this cohort that were successfully sequenced by the MFT NGS custom panel to be 385, with 76 unsuccessful and an overall success rate of 84%.

**Table 1.** Summary of success rate and regional variability by cohort. Each cohort displayed a range of success rates and regional variability amongst referrers.

|  | Northwest Lung | Wythenshawe Lung | Ovary |
| --- | --- | --- | --- |
| Time Period For Sample Collection (Months) | 6 | 17 | 17 |
| Number of Referring Centres | 23 | N/A |  |
| Total Number Samples | 801 | 461 | 882 |
| Number Successful | 591 | 385 | 750 |
| Number Unsuccessful | 210 | 76 | 132 |
| Success Rate (%) | 74 | 84 | 85 |
| Top 5 Volume Referring Centre Sample Contribution (%) | 61 | N/A | 28 |
| Top 5 Volume Referring Centre Success Rate Range (%) | 68 - 81 | N/A | 81 – 84 |

Considering Northwest Ovarian samples, analysis of the overall cohort of 882 ovarian NGS samples processed at the NWGLH over a 17-month time period from 01/01/2021 to 01/06/2022, revealed the overall number of samples within this cohort that were successfully sequenced by the MFT NGS custom panel to be 750, with 132 unsuccessful and an overall success rate of 85%. 85 different individual referring hospitals were identified from the sample cohort, with 122 samples (13.8%) classified as unknown from the available data, leaving n=760. Whilst there were significant regional variations in the number of samples referred for sequencing, overall success rates varied less than lung samples amongst individual referrers, ranging from 81% - 84% amongst the top five high-volume referrers (Table 1.).

### b) Multivariate Logistic Regression Analysis of Pre-Analytic Factors

For multivariate analysis of the Northwest Lung Cohort, 1 sample was excluded from the analysis as a result of unclear data (Table 2.). A sample type of image-guided biopsy (both CT and US) (Odds Ratio: OR=2.21, [1.06; 4.65], p= 0.0355) was associated with higher rates of unsuccessful QIAseq Status amongst samples run. Low concentration of extracted sample (OR=0.92, [0.9; 0.94], p <0.0001) was also significantly associated with higher rates of unsuccessful QIAseq Status amongst samples from the cohort analysed. Nanodrop 280/260 Ratio (OR=0.98, [0.92; 1.04], p= 0.4576), Nanodrop 230 /260 Ratio (OR=1.0, [0.99; 1.0], p= 0.5942) for DNA purity with both methods being susceptible to small changes in pH and contaminants, Macrodissection = No (OR=0.967 [0.912;1.12], p=0.234), NCC Content = <20% (OR=1.43 [1.010;1.850], p= 0.092), Extraction material = slide (OR=1.15 [0.865;1.435], p=0.547), Extraction material = wax (OR=1.32 [1.147;1.493] p=0.289), Sample Type = Biopsy (OR=1.27, [0.78; 2.07], p= 0.338), Sample Type = Pleural biopsy/aspirate (OR=0.52, [0.22; 1.23], p= 0.1372), Sample Type = Peripheral biopsy/aspirate (OR=0.78, [0.27; 2.2], p= 0.6352) and Sample Type = EBUS (OR=0.9, [0.52; 1.56], p= 0.7091), were all not associated with the rate of QIAseq Status = Unsuccessful.

**Table 2.** Multivariate logistic regression analysis table of pre-analytic factors related to NGS QIAseq success.

|  | Variable | Modality | Odds of Failure | p-value |
| --- | --- | --- | --- | --- |
| **Lung North West** | Intercept |  | 2.23 [1.3;3.83] | 0.00375* |
|  | Concentration |  | 0.918<br>[0.902;0.935] | 5.27e-20* |
|  | Nanodrop 280/260 Ratio |  | 0.977<br>[0.92;1.04] | 0.458 |
|  | Nanodrop 230/260 Ratio |  | 0.998<br>[0.993;1.0] | 0.594 |
|  | Macrodissection | Yes (reference) |  |  |
|  |  | No | 0.967<br>[0.912;1.12] | 0.234 |
|  | Extraction Material | Shavings (reference) |  |  |
|  |  | Slide | 1.15<br>[0.865;1.435] | 0.547 |
|  |  | Block | 1.32<br>[1.147;1.493] | 0.289 |
|  | NCC Content | >20% (reference) |  |  |
|  |  | <20% | 1.43<br>[1.010;1.850] | 0.092 |
|  | Sample Type | Excision<br>(reference) |  |  |
|  |  | Biopsy | 1.27<br>[0.779;2.07] | 0.338 |
|  |  | EBUS | 0.901<br>[0.522;1.56] | 0.709 |
|  |  | Peripheral<br>biopsy/aspirate | 0.778<br>[0.275;2.2] | 0.635 |
|  |  | Pleural<br>biopsy/aspirate | 0.518<br>[0.218;1.23] | 0.137 |
|  |  | Radiological guided<br>Biopsy (CT/US) | 2.21 [1.06;4.65] | 0.0355* |
| Lung Wythenshawe | Macrodissection | No (reference) |  |  |
|  |  | Yes | 1.19 [0.53;2.69] | 0.669 |
|  | Sample type | Excision<br>(reference) |  |  |
|  |  | Biopsy | 0.776<br>[0.266;2.26] | 0.642 |
|  |  | EBUS | 1.42 [0.489;4.1] | 0.522 |
|  |  | Peripheral<br>Biopsy/aspirate | 2.14<br>[0.501;9.13] | 0.304 |
|  |  | Pleural<br>Biopsy/aspirate | 1.18<br>[0.292;4.79] | 0.815 |
| Ovary Northwest | Intercept |  | 5.14<br>[0.524;50.4] | 0.16 |
|  | Concentration |  | 0.932<br>[0.897;0.968] | *0.000312 |
|  | Nanodrop 260:280<br>Ratio |  | 0.307<br>[0.151;0.624] | *0.00111 |
|  | Nanodrop 260:230<br>Ratio |  | 0.528<br>[0.323;0.863] | *0.0109 |
|  | NCC Content | <20% (reference) |  |  |
|  |  | >20% | 0.885<br>[0.373;2.1] | 0.781 |
|  |  | >50% | 0.956<br>[0.279;3.28] | 0.944 |
|  | Macrodissection |  | 2.03 [1.09;3.79] | 0.0254* |
|  | Sample type | Excision<br>(reference) |  |  |
|  |  | Biopsy | 0.776<br>[0.266;2.26] | 0.642 |
|  |  | Peripheral<br>Biopsy/aspirate | 2.14<br>[0.501;9.13] | 0.304 |

Furthermore, multivariate analysis of the Wythenshawe hospital cohort (Table 2.) revealed, DNA concentration (OR=0.93, [0.9; 0.97] and DNA quality assessed by Nanodrop 260:280 Ratio (OR=0.31, [0.15; 0.62], p= 0.0011) and Nanodrop 260:230 Ratio (OR=0.53, [0.32; 0.86], p= 0.0109), p= 0.0003) were associated with lower rates of unsuccessful NGS sequencing. NCC Content = >20% (OR=0.88, [0.37; 2.1], p= 0.7808), NCC Content = >50% (OR=0.96, [0.28; 3.28], p= 0.9435), Macrodissected (OR=1.19, [0.53; 2.69], p= 0.6695), Sample type = Biopsy (OR=0.776 [0.266;2.26] p=0.642), EBUS (OR=1.42 [0.489;4.1] p=0.522), Peripheral Biopsy/aspirate (OR=2.14 [0.501;9.13] p=0.304) and Pleural biopsy/aspirate (OR=1.18 [0.292;4.79] p=0.815) were found not to be associated with the unsuccessful rate of NGS sequencing.

Considering multivariate analysis of the Northwest Ovarian cohort (Table 2.), the presence of macrodissection (OR=2.03, [1.09; 3.79], p= 0.0254) was associated with higher rates of unsuccessful NGS, whilst both higher DNA concentration (ng/ul) (OR=0.83, [0.79; 0.87], p <0.0001) and higher DNA quality measured by Nanodrop 260:280 (OR=0.24, [0.11; 0.55], p= 0.0007), but not Nanodrop 260:230 (OR=1.01, [0.8; 1.28], p= 0.9229), were associated with lower rates of unsuccessful NGS. Although neither neoplastic cell content = >20% (OR=0.74, [0.33; 1.63], p= 0.4535), or neoplastic cell content = >50% (OR=0.5, [0.23; 1.11], p= 0.0888) was associated with the rate unsuccessful NGS a trend approaching significance with increasing cell count was observed.

### c) Univariate Analysis of Influence of Macrodissection on Success Rate

Analysis of the Northwest Lung Cohort for differences in NGS success rate as a result of macrodissection revealed trends with marginally higher success rates (74.1% v 70.5%) in non-macrodissected samples; however, this did not reach significance on univariate analysis. Similarly, within the Wythenshawe hospital cohort success rates for macrodissected samples were 76% (38/50) compared to 84.4% (347/411). Although a trend towards better success rates without macrodissection was observed, similar to the overall Lung NGS cohort, the difference did not reach statistical significance. However, within the Ovarian sample cohort, the absence of macrodissection was found to be associated with a greater likelihood of NGS success (78% v 87.4%, p=0.001 Chi-squared test) (Fig. 1.), likely to reflect original sample insufficiency which has a significant impact on NGS success as revealed by earlier multivariate analysis.

**Figure 1.**
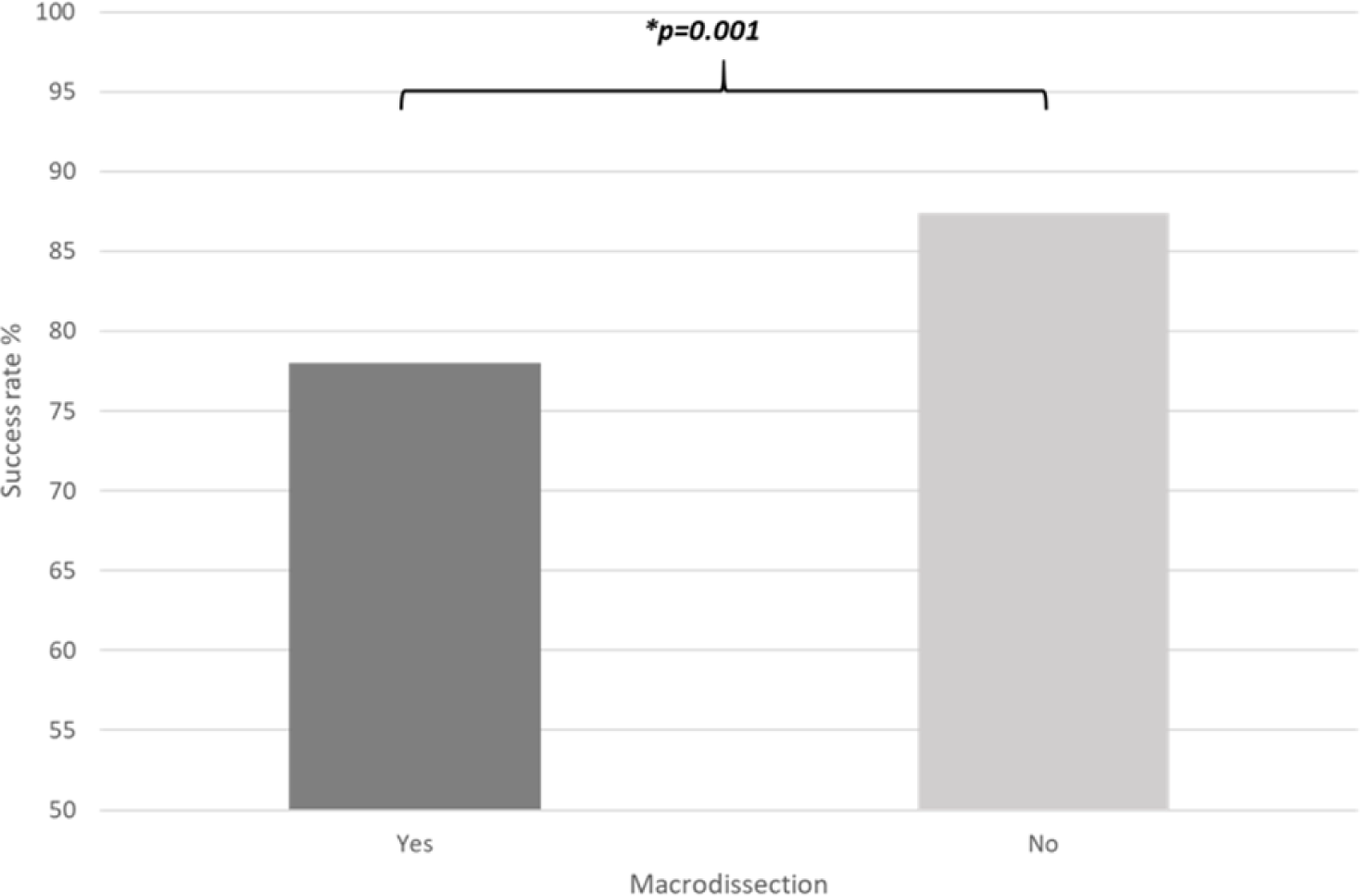
Ovarian NGS success rate according to sample macrodissection. A higher success rate in non-macrodissected samples was observed (p=0.001, Chi-squared test).

### d) Univariate Analysis of Lung Sample Type on Success Rate

NGS success was found to be strongly influenced by sample type within the Wythenshawe lung cohort, with the overall success rate of surgical excision samples 98.7% (160/162) whilst only 75.3% (225/299) amongst non-surgical excision samples (OR = 0.038; CI[0.0092; 0.16]; p<0.001) (Fig. 2.). Further detailed analysis of sample type revealed a wide variation in types of sample other than the most common type, surgical excision specimens (156/461, 33.8%), with EBUS-FNA samples (107/461, 23.2%) and Lung Biopsy samples (87/461, 18.9%) being the next most common. NGS success was strongly influenced by specific sample type on univariate analysis with the success rate of surgical excision samples ∼100% whilst only 71.9% for EBUS samples and 74.7% for lung biopsy samples within the dataset (p<0.001 Chi-squared test).

**Figure 2.**
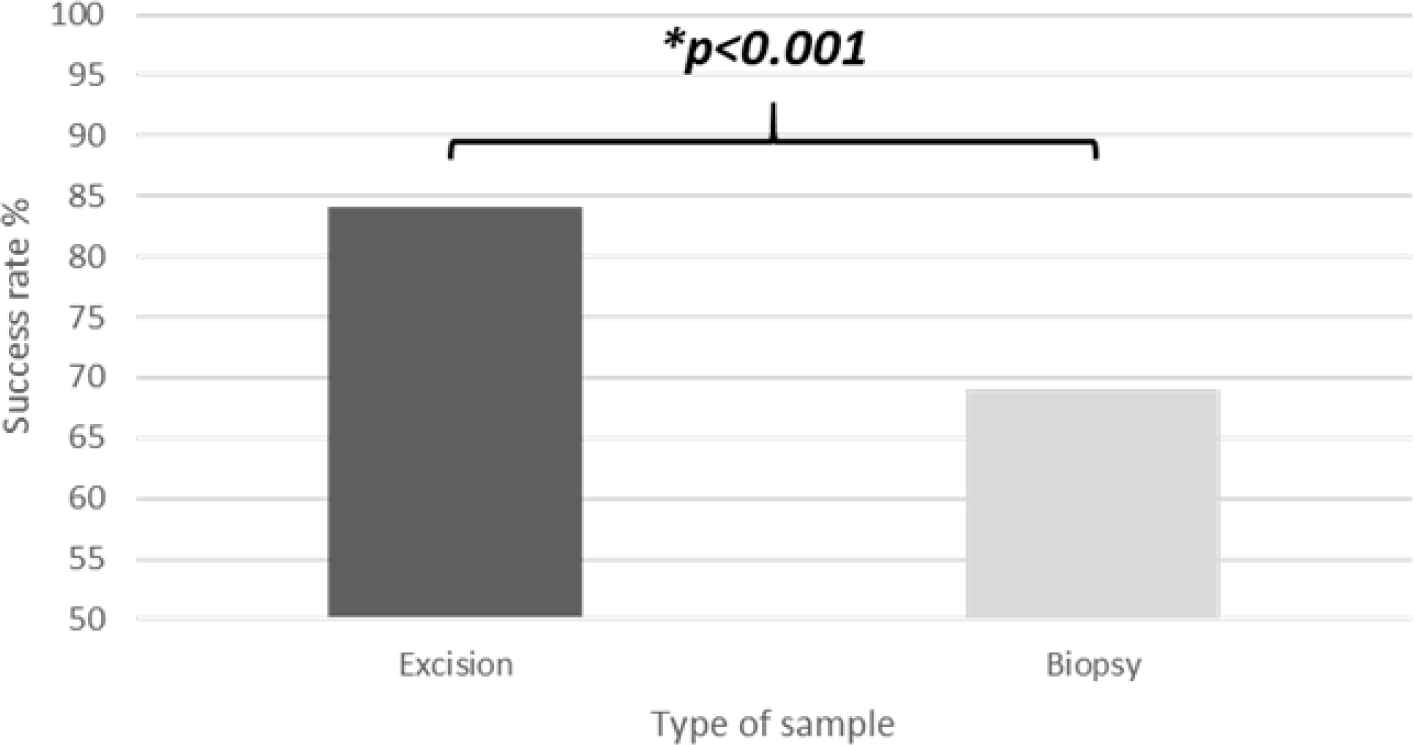
Lung NGS success rates (Wythenshawe) in relation to surgical or non-surgical excision. Biopsy (non-surgical excision) samples have a significantly lower success rate compared to surgical excision samples within the cohort that have ∼100% success rate (p<0.001, Chi-squared).

**Figure 3.**
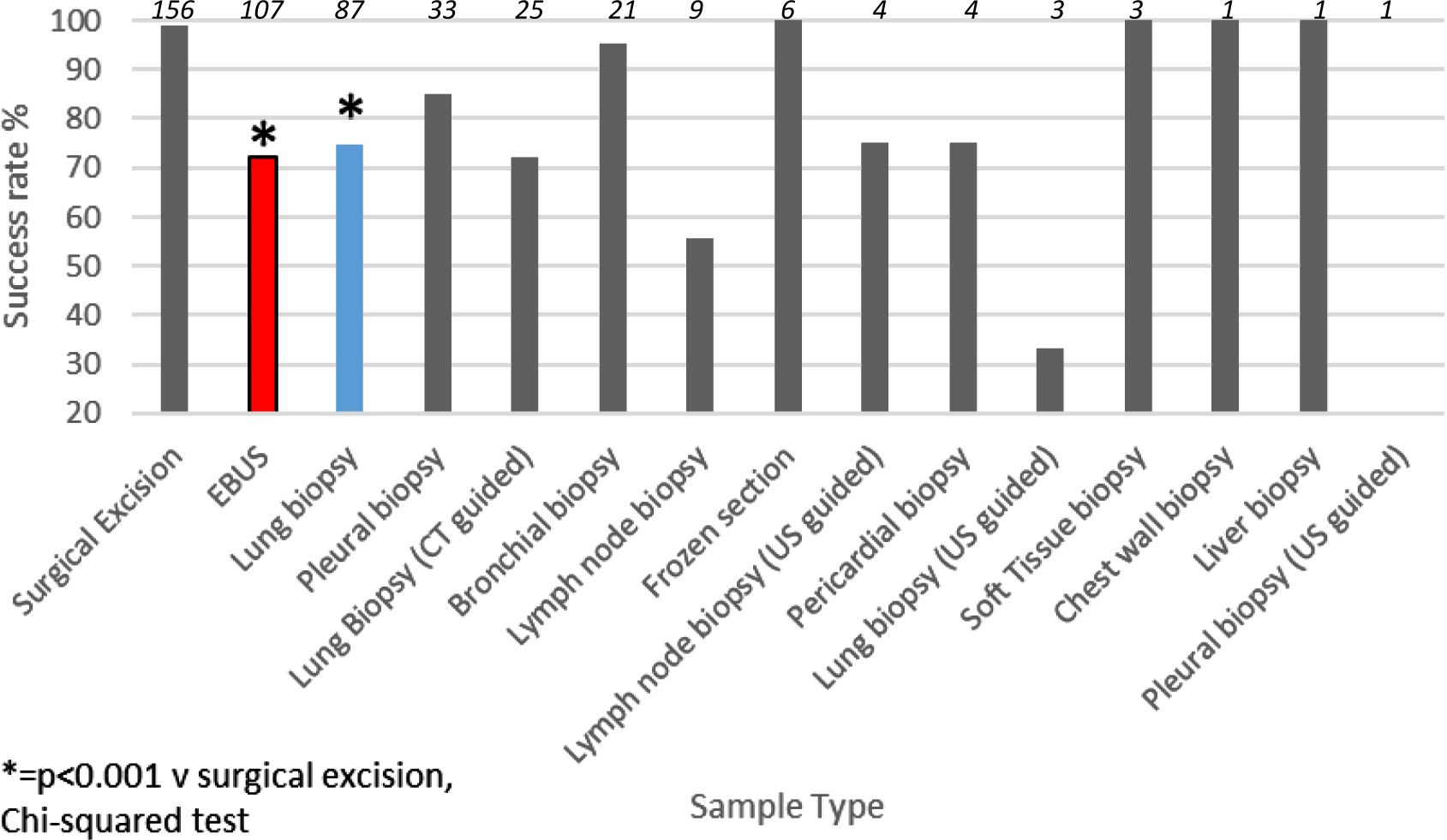
Lung NGS (Wythenshawe) success rate according to specific sample type. Sample type ordered by frequency in dataset, most common to left. Compared to surgical excision samples that have ∼100% NGS success rate, EBUS and Lung Biopsy samples, whilst being the next most common sample type, have significantly lower success rates at 71.9% (red) and 74.7% (blue) respectively (the 95% CI was wide for the rarer specimen types). Numbers above bars indicate total numbers of samples analysed.

Following this analysis, as lung biopsy samples and EBUS samples were found to be significantly poorer performing, further pre-analytic data were then gathered in a targeted way to assess whether the number and nature of core biopsies for lung biopsy samples and the site of harvest and fixation time for EBUS samples, had any association with NGS success rates.

### e) Univariate Analysis of Lung Biopsy Characteristics on Success Rate

Further targeted analysis of Wythenshawe cohort lung biopsy samples with regards the number of core biopsies provided to the pathology lab after sample acquisition (i.e. providing greater choice for best core selection) revealed a marked graded difference in success rate dependent on the number of cores taken with ≤2 cores associated with a 66.6% success rate, 3 cores displaying 76.2% and ≥ 4 cores displaying 81.8%. Median values of number of cores were respectively 2.51 (IQR 1.0) and 2.0 (IQR 2.0) in patients for which NGS status was successful and unsuccessful respectively (p=0.099). Furthermore, the median values of length of longest core (mm) were respectively 13.5 (IQR 7.0) and 12.0 (IQR 9.0) in patients for which NGS status was successful and unsuccessful respectively (p=0.079).

Analysis of whether site from which EBUS-FNA samples were obtained had an influence on NGS success rates was performed (Table 3.). Anatomical sites that were more difficult to sample: paratracheal regions (predominantly lymph node stations 2 and 4), carinal (predominantly lymph node stations 7 and 10) compared to easier to sample sites: hilar (predominantly lymph node stations 11 and 12) and lung, revealed significantly higher success rates for hilar and lung regions (90.3% and 100% respectively) compared to paratracheal and carinal regions (68.3% and 67.5% respectively) (*p=0.0086, Chi-squared test*).

**Table 3.** Summary of success rate according to anatomical level of EBUS-FNA. Significant differences in success rates (p<0.05, Chi-squared test) exist between sampling for paratracheal and carinal regions compared to hilar and lung regions (Lymph node stations shown in brackets).

| EBUS Site | Number of samples | Successful | Unsuccessful | %Success |
| --- | --- | --- | --- | --- |
| Paratracheal (2,4) | 60 | 41 | 19 | 68.3 |
| Carinal (7,10) | 37 | 25 | 12 | 67.5 |
| Hilar (11, 12) | 31 | 28 | 3 | 90.3 |
| Lung | 9 | 9 | 0 | 100 |

### f) Univariate Analysis of Influence of Fixation Time on Success Rate

Data with regard to fixation time within the Wythenshawe hospital cohort related to the EBUS-FNA cases included in our sample cohort (n=107) was further analysed. Two cases were excluded due to lack of data. Only 49.5% of samples (52/105) met the existing local guideline for maintaining fixation time <24 hours, with 62% (32/52) of prolonged fixation samples being received in the lab the following day. Although statistical significance was not observed with increasing time of sample reception (days to receive 0 i.e. same day processing success rate, 73.9% v days to receive 1 at 65.6%) as well as days to process the sample (days 1-4 ranging from 78.8% to 47%) a clear trend in decreasing success rates with increasing time was observed. On grouping the samples and applying a cut-off at 48 hours (allowing for unavoidable delays in reception or processing of samples) a statistically significant decrease in success rate (p=0.016 on Chi-squared test) was revealed.

The availability of fixation data for ovarian samples was limited precluding further analysis.

## Discussion

The pre-analytic phase of next-generation sequencing (NGS) analysis is influenced by multiple factors that can impact the success of the workflow. This study aimed to investigate the pre-analytic steps of sample acquisition and processing that can affect the NGS pipeline. Efforts to optimize methods for NGS testing using (FFPE) samples have resulted in improved success rates within clinical laboratories. Dropout rates as high as 46% due to poor DNA quality or low DNA yield following extraction have been reported, but recent studies, including this one, have shown higher success rates for lung and ovarian cancer NGS (28). For instance, success rates of 74% for lung cancer and 85% for ovarian cancer have been reported, aligning with similar findings in the literature (29, 30). However, some institutions have reported even higher success rates, such as a South Korean study that achieved an overall success rate of 95.9% for NGS analysis of 1083 FFPE samples (31).

Specific studies focusing on lung cancer have demonstrated improvements in success rates by increasing the number and size of biopsy samples, particularly in cases where collaboration between clinicians and pathologists during sample collection was enhanced (32). Similarly, modifications to routine DNA extraction protocols, such as the addition of the enzyme Uracil-N-Glycosylase, have been shown to reduce formalin fixation-related artifacts and improve NGS success rates to nearly 100% in ovarian cancer testing (33). The variation in success rates observed across different referring pathology laboratories in this study (ranging from 46% to 100% for lung cancer and 33% to 100% for ovarian cancer) may be attributed to both the volume of referrals and differences in pre-analytic practices and standardization among regional pathology laboratories.

Consistent with previous studies, DNA yield was identified as the most significant factor associated with NGS success across all datasets and analyses in this study (34). Comparison of different DNA extraction methods for large-scale genomic analysis demonstrated the superiority of modified silica-based commercial kits over traditional methods (35). The cobas® kit used for DNA extraction from FFPE specimens at NWGLH showed high yields of good quality DNA. However, as this study did not assess variations in specific extraction steps and protocol adherence, further investigation is needed to determine the impact of these factors on overall DNA yield potentially through approaches assessing paired data from COBAS and Qiasymphony extraction method. Nevertheless, improvements in tissue volume and fixation, which were more readily assessed, were shown to have a significant influence on NGS success rates.

The type of specimen plays a crucial role in NGS success, as demonstrated by the higher success rates in lung surgical excision samples compared to biopsy samples, particularly EBUS-FNA and radiologically guided lung biopsies, where tissue availability is limited (32). This disparity is further evident in the higher overall success rates observed for ovarian NGS, which includes a larger proportion of surgical excision samples compared to lung samples. Additionally, the number and size of lung core biopsy samples were found to significantly influence NGS success rates, highlighting the importance of initial tissue volume. Notably, the existing local guidelines do not provide recommendations specifically addressing these factors.

Pooling shavings from multiple blocks to maximize tissue for DNA extraction should be considered, as distributing cores across multiple blocks may result in underutilization of available tissue. Moreover, using a wider gauge core biopsy needle has been shown to improve NGS success rates in EBUS lung biopsy, in contrast to the finer 22 needle used in previous studies (29). Interestingly, CT-guided biopsy samples showed lower success rates in the overall lung dataset, although they generally provide larger tissue amounts. However, the sample size for CT-guided biopsies was small, indicating a need for further investigation and reflex testing of all adenocarcinomas is likely to result in increased numbers for future analysis.

The anatomical location of samples collected through the EBUS approach for lung biopsy can also impact NGS success rates. Paratracheal and carinal samples demonstrated significantly lower success rates compared to hilar and lung regions. This finding aligns with previous research indicating reduced diagnostic accuracy in certain nodal stations within the paratracheal and carinal regions (36). On the basis of our findings it would be important to recommend to clinicians acquiring EBUS specimens that they should preferably obtain specimens from the primary tumour, even when lymph nodes are positive.

Achieving a high neoplastic cell count (NCC) is crucial to reduce the risk of false negative NGS results. This study found a significant association between NGS success and NCC content greater than 20% in lung samples and a graded manner for ovarian samples establishing NGS success to be correlated with a better sample of tumour. However, determining NCC remains subjective, and there is substantial variation in practice as evidenced by a recent Europe-wide study (37). Consensus among experts suggests that pathologists should determine NCC content based on the area of the specimen with the lowest density of inflammatory cells but the highest density of neoplastic cells following dissection of specimens (38).

Macrodissection, a challenging process involving the visual identification and tracing of tumour cellson FFPE slides, showed higher NGS failure rates in both lung and ovarian cancer datasets. This is likely to reflect the fact that the need for macrodissection is directly associated with poor initial sample provision; however, technical drawbacks in macrodissection methods may also contribute with a lack of standardization and objectivity in the process. Other approaches, such as microdissection or laser microdissection, offer the potential for improved tissue areas for DNA extraction, resulting in higher NGS variantdetection rates (39). However, optimization of these technologies within the NHS is still needed and initial studies assessing samples with good tumour percentage, in which microdissection is not beneficial, analysed with and without microdissection, would be first necesssary to assess whether it is detrimental.

Prolonged tissue fixation beyond 48 hours had a significantly detrimental effect on NGS success rates for lung EBUS samples. Concerningly, only around 50% of EBUS samples from a high-volume lung biopsy referral centre met the existing guidelines for fixation times of less than 24 hours. This indicates that there may be a significant benefit from adopting an extended working day so that pathology laboratories can receive and process specimens on the day that they are taken and also consider Saturday embedding to avoid the maintenance of specimens in formalin over the weekend. Formalin, while widely used as a fixative, can negatively impact DNA integrity by causing fragmentation and nucleotide transitions (40). The present study used CytoRich Red for fixation, which is superior to formalin for cytology specimens but still has drawbacks related to fixation times. Optimal fixation times of 12 to 24 hours in buffered formalin at room temperature have been suggested for DNA extraction and subsequent genomic analysis (40). These results indicate that a significant number of lung specimens are processed with inappropriately long fixation times, which can affect efficient DNA analysis.

Conversely, longer fixation times (∼2.5 days) were associated with successful NGS and higher DNA yields in ovarian samples within the Manchester dataset. The Royal Preston Hospital achieved a mean fixation time of 4 days with a 100% NGS success rate. Larger specimens may require longer fixation times for formalin to adequately penetrate and preserve tissues, in line with the existing local guidelines (27), however, specific evidence providing differences in length of optimal fixation according to type and size of sample remain lacking. Further analysis of ovarian samples is needed to determine optimal fixation times, but the data suggest that guidelines should be updated to consider this specific aspect in ovarian cancer samples.

Future studies should focus on identifying the reasons for failures in adhering to regional recommendations and pinpointing critical delays in the sample processing pathway that impact fixation time. Adherence to improved sample acquisition methods, especially for specific techniques and anatomical sites, alongside reductions in fixation times, may help achieve higher NGS success rates. Collaborative efforts between NHS genomic hubs and pathology laboratories, along with standardized protocols and updated guidelines, may further improve outcomes in the future.

## Data Availability

All data produced in the present study are available upon reasonable request to the authors.

## Acknowledgements including a statement defining funding sources

The authors would like to acknowledge all referring pathology laboratories sending samples for analysis to the NWGLH, alongside all Manchester Genomics laboratory genetic technologists and bioinformaticians involved in sample processing and data analysis for this study. Special acknowledgement should be made of Henry Pickup and Deborah Lakeland of Lancashire Teaching Hospitals NHS Trust Cellular Sciences and Genomic Services who provided specific information regarding Ovarian sample fixation time for this study. No separate funding was obtained for this study which was completed as part of a National School of Healthcare Science Master’s Research Project utilising readily available local resources and collaborations.

## Statement of author contributions

PBh, GJB and HS all participated in designing the study. DG and SR provided data from the Manchester Genomics Laboratory, whilst PBi and BE provided data from Wythenshawe Hospital for analysis as part of the study. PBh and PBi performed data analysis, whilst PBh, PBi, GB and HS were all involved in the writing editing and approval of the finished manuscript.

---

**Figure 3.**
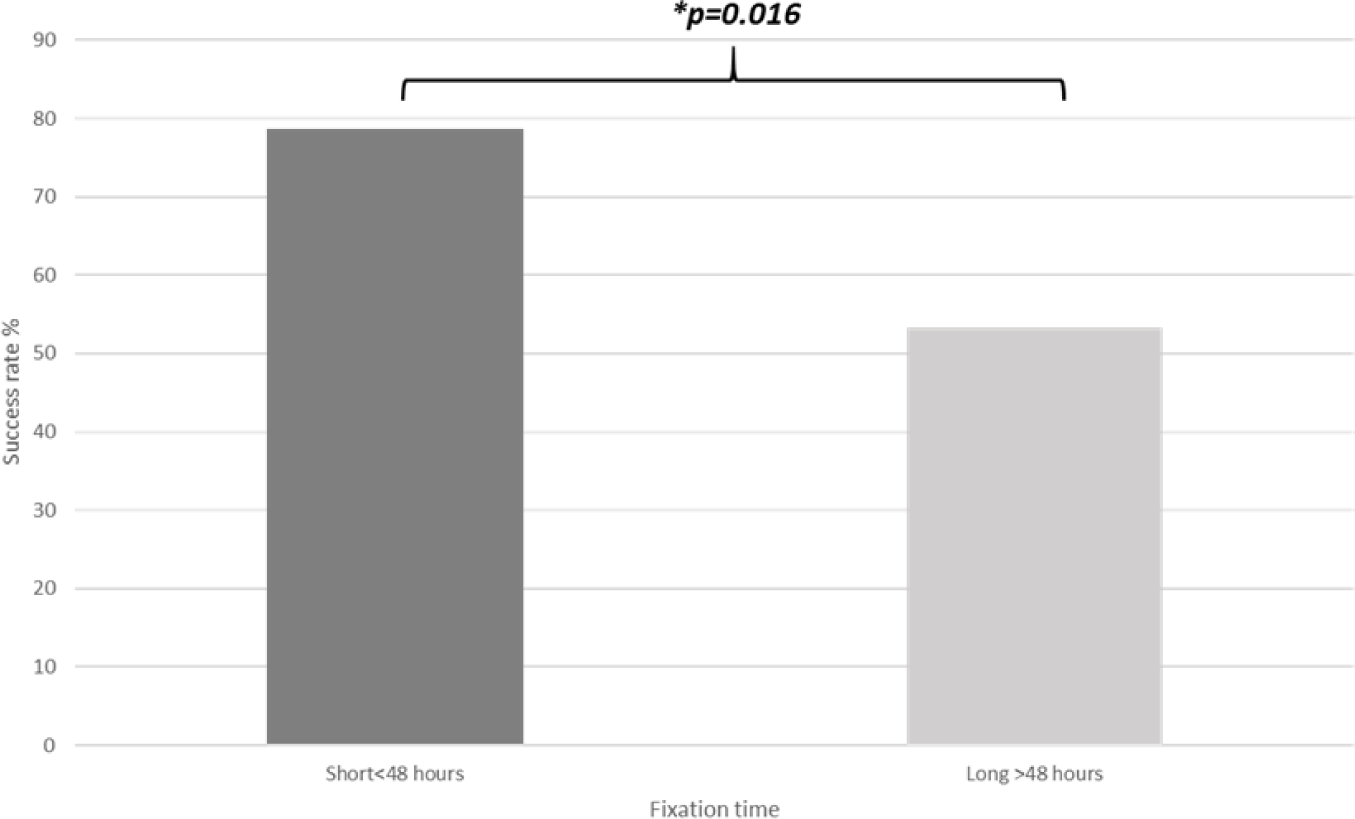
Effect of >48 hours fixation time on EBUS-FNA NGS success rate (Wythenshawe samples). NGS success rates was significantly lower (53.3%v78.7%) after 48 hours fixation time (p=0.016, Chi-squared test).

